# A Fast, Lightweight, and Generalizable Deep Neural Network for the Detection of Atrial Fibrillation

**DOI:** 10.1101/2025.11.13.25340129

**Authors:** Harshit Mishra, Farhan Adam Mukadam, Nachiket Makwana, Pradyot Tiwari, K. Subramani, K. V. S. Hari

## Abstract

Atrial fibrillation (AFib) represents a critical diagnostic challenge in clinical cardiology, calling for automated detection systems capable of robust performance across diverse clinical environments. We present a computationally efficient deep neural network architecture for AFib detection that demonstrates exceptional generalizability despite training on a modest dataset. Our convolutional neural network, comprising 17 million parameters, was trained on 67,432 12-lead electrocardiograms and subsequently validated on over 1.1 million ECG recordings spanning six independent public datasets. On CPU, the model processes a 10-second ECG in 200 ms, enabling real-time inference capabilities.

Our model achieves state-of-the-art performance, with an average area under the receiver operating characteristic curve (AUROC) of 0.97, an average sensitivity of 0.83, and an average specificity of 0.96 across six external validation cohorts. These metrics rank among the highest reported in the literature, while preserving computational efficiency suitable for resource-constrained environments.

A key innovation of our approach is the implementation of channel-masking methodology, enabling seamless operation across variable lead configurations without model retraining. This flexibility allows deployment from single-lead ambulatory monitors to comprehensive 12-lead clinical systems using identical network weights. Gradient-based saliency analysis confirms the model’s attention to physiologically relevant features, particularly P-wave morphology and lead II characteristics, thereby enhancing clinical interpretability and trustworthiness.

Our findings also establish that a single, well-curated small training dataset can yield a compact yet highly generalizable AFib detection system suitable for deployment across diverse clinical settings, from critical care monitoring to ambulatory screening applications. The combination of robust cross-dataset performance, computational efficiency, and clinical interpretability positions this approach as a viable solution for largescale AFib diagnosis and monitoring programs.

## 1. Introduction

Atrial fibrillation (AFib) is the most common sustained cardiac arrhythmia. Global prevalence already exceeds 33.5 million persons and is projected to reach 17.9 million in Europe and 12.1 million in the USA by 2030 [1, 2]. Each year AFib accounts for nearly 454,000 US hospitalizations and 158,000 deaths, and adds up to $26 billion to direct health-care spending [4, 3]. In critical care environments the problem is magnified: new-onset AFib occurs in 5–15% of intensive-care admissions and independently raises 30-day mortality by 40–60% [5, 6]. AFib also confers a five-fold stroke risk, triples heart-failure incidence, and doubles all-cause mortality [7]. Contemporary guidelines therefore emphasize early rhythm identification to guide anticoagulation and rhythm control strategies [8, 9]. Yet manual ECG interpretation is resource intensive and error prone, motivating research into fully automated detectors deployable at the bedside or on wearables.

### Evolution of automated AFib detection

First-generation algorithms exploited heart rate variability: Tateno & Glass [10] achieved 94.4% sensitivity and 97.2% specificity from RR-interval histograms, while Dash *et al*. combined time- and frequency-domain descriptors to surpass 98% accuracy [11] on their internal test sets. Subsequent hand-crafted entropy measures [12] improved robustness but required expert-driven feature engineering. All these handcrafted approaches required extensive domain expertise for feature engineering and were prone to failure under adverse or unseen conditions

### Deep-learning era

Rajpurkar *et al*. framed arrhythmia detection as a sequence-to-sequence task and ran raw, single-lead ECG through a 34-layer 1-D ResNet—16 [13]. Attia *et al*.’s “AI-ECG” preserves the purely convolutional philosophy but adapts it to 12 leads: stacked 1-D convolutional layers with progressively wider receptive fields [14]. Ribeiro *et al*. port the 2-D image ResNet to one-dimensional signals for multi-label diagnosis on 12-lead input—residual that outputs six common abnormalities [15]. Moving beyond pure CNNs, Natarajan *et al*.’s “wide-and-deep Transformer” first embeds the waveform with a small convolutional front-end, adds positional encodings, then passes the sequence through an eight-layer Transformer encoder (multi-head self-attention, *d* = 256) whose output is concatenated with 22 handcrafted “wide” ECG features before the final classifier [17]. However,most studies are evaluated on a single site; true cross-cohort generalization and deployability remain open challenges.

### Our contribution

We present a lightweight and fast 17M-parameter CNN, trained on a small but carefully curated HDXML corpus—specifically, the Heart Disease HDXML Strain dataset by Tse et al. [18]—comprising 67,432 12-lead ECGs. We externally validated on more than 1.1 million ECGs across six public datasets including MIMIC-IV critical-care recordings. Built-in channel attention and dual-path fusion permit identical weights to process anything from a single-lead wearable trace to a full 12-lead ECG, meeting the practical needs of both outpatient monitors and ICU telemetry systems.

## 2. Methodology

### 2.1. Dataset Preparation

We pooled eight open-access 12-lead ECG datasets totaling 1.37 million tracings (see Table 1): HDXML [18] (hereafter referred to as HDXML), CODE-15 [19], Chapman–Shaoxing [20], Chapman-10k [21], MedalCare-XL [22], PhysioNet 2020 (Offline-TS) [23], PhysioNet CinC 2021 [24], and the MIMIC-IV ECG dataset [25]. Each dataset is loaded by a dedicated dataloader that harmonizes file formats and maps native rhythm labels to our binary AFib/non-AFib schema. All signals, regardless of origin, were processed as follows:

**Table 1:** Sample distribution across all datasets (AFib vs Non-AFib)

| Dataset | AFib Samples | Non-AFib Samples | Total Samples |
| --- | --- | --- | --- |
| code 15 dataset | 3,866 | 188,202 | **192,068** |
| chapman train dataset | 502 | 20,114 | **20,616** |
| chapman 10k train dataset | 1,441 | 7,075 | **8,516** |
| offline ts train dataset | 294 | 5,306 | **5,600** |
| physionet 2021 train dataset | 4,223 | 66,379 | **70,602** |
| MIMIC-IV test set | 78,417 | 721,617 | **800,034** |
| HDXML Dataset | 10,471 | 56,961 | **67,432** |

- **Resampling –** all recordings were resampled (or kept) at 500 Hz.
- **Min–max normalisation –** each lead was linearly scaled to the range [−1, 1].
- **Fixed-length framing –** signals were zero-padded or truncated to 5000 samples (≈10 s) to enable mini-batch training.

Augmentations (training splits only): A single stochastic transformation was sampled per mini-batch from the list below; all augmentations preserve class membership.

- **Temporal transforms:** random circular phase shift; variable-length leading/trailing zero shifts.
- **Morphological noise:** baseline drift (linear), baseline wander (low-frequency sinusoids), EMG-like muscle artefacts (20–100 Hz band-passed Gaussian noise).
- **Channel manipulations:** accidental lead reversal, random lead swapping, electrode contact artefacts (spikes, drop-outs, steps, jitter).
- **Spectral smoothing:** FFT low-pass (*f*_c_ = 40 Hz, Hann transition) to simulate analogue antialias filtering.
- **Spatial masking:** per-sample binary lead mask to mimic missing channels (used later by the interpretability head).

All transformations were implemented on-the-fly to avoid duplicating data on disk and were disabled for validation, internal testing, and all external evaluations.

### 2.2. Model Architecture

We developed a deep learning architecture for automated Atrial Fibrillation (AFib) detection from 12-lead ECG signals. The architecture leverages the power of convolutional neural networks (CNNs) to extract and analyze temporal and spatial features from ECG signals. The proposed ECG classification model consists of three main blocks: Feature Extraction Network, Dense Feature Extraction, and Classification Head. Each block is designed to progressively extract and refine the features of the raw ECG signals for accurate binary classification, as illustrated in Figure 1.

**Figure 1:**
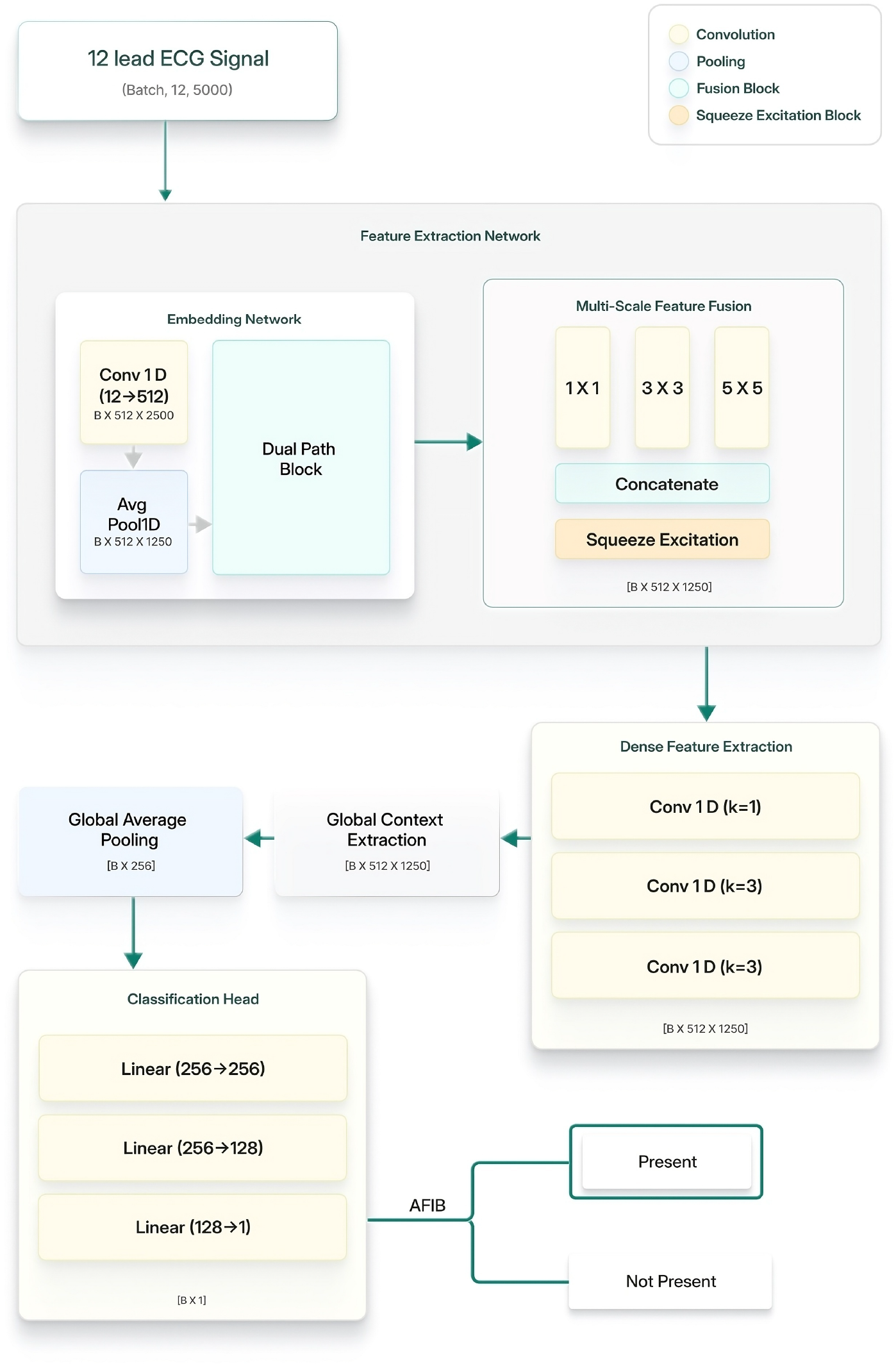
Overall model architecture showing the three main components: embedding network, feature extraction network, and classification head for AFib detection from 12-lead ECG signals.

#### 2.2.1. Feature Extraction Network

The feature extraction network serves as the primary feature learning component, transforming raw ECG signals into rich, multi-scale representations through two complementary sub-modules: Embedding Network and Multi-Scale Feature Fusion.

##### Embedding Network

The embedding network transforms raw ECG signals into compact feature representations. Once the 12×5000 signal is loaded, an initial 1-D convolution with 7×7 kernel, stride 2, and padding 3 is applied to generate a 512×2500 dimensional output. This is followed by batch normalization and ReLU activation to ensure stable training and introduce non-linearity for effective feature extraction. An average pooling layer with 3×3 kernel size, stride 2, and padding 1 then downsamples the output to 512×1250 dimensions.

The core component is the dual path block, which enhances feature extraction through a custom neural network comprising three parts: 1) A residual path, 2) A dense connection path, and 3) Squeeze and Excitation Mechanism. The residual path consists of two stacked convolutional layers with large receptive fields using 15×15 convolutional kernels, enabling capture of long-range temporal dependencies. The dense connection path utilizes 1×1 convolutions for efficient feature transformations.

The Squeeze-and-Excitation (SE) block operates as a channel-wise attention mechanism designed to enhance the representational power of convolutional neural networks by adaptively recalibrating channel-wise feature responses. The SE block seamlessly integrates with existing architectures and improves feature selectivity and overall network efficiency through adaptive channel weighting.

##### Multi-Scale Feature Fusion

The multi-scale feature fusion module captures contextual information at different granularities within the ECG signal processing pipeline. The module utilizes three distinct parallel convolutional branches with kernel sizes of 1×1, 3×3, and 5×5, each applied to the input feature maps. These convolutions capture different levels of feature granularity: the 1×1 convolution captures point-wise information and cross-channel interactions, the 3×3 convolution captures local contextual features and short-term temporal patterns, and the 5×5 convolution extracts broader contextual patterns and longer-term temporal dependencies. The outputs are concatenated and processed through a squeeze-and-excitation mechanism before being fused back to the original channel dimension with a residual connection.

#### 2.2.2. Dense Feature Extraction

The dense feature extraction block refines and transforms the multi-scale features from the feature extraction network through two sequential processing stages.

##### Convolution

After the initial feature extraction, the dense feature extraction process begins with a sequence of convolutional layers aimed at progressively refining and transforming the initial embeddings. The process starts with batch normalization applied to the embeddings, followed by a series of convolutional layers:

- Initial 1×1 convolutional layer reduces the dimensionality to 512 channels
- Two sequential 3×3 convolutional layers with padding capture local temporal patterns
- Batch normalization and ReLU activation are applied after each convolution
- Final batch normalization ensures stable feature distributions

##### Global Context Extraction

The global context extraction module transforms the refined convolutional features into a compact global representation. A 1×1 convolution reduces the feature dimensionality to 256 channels, followed by batch normalization and ReLU activation. A squeeze-and-excitation block is then applied for final feature refinement and adaptive channel weighting. This stage prepares the features for the final classification by capturing global temporal patterns and contextual information across the entire ECG sequence.

#### 2.2.3. Classification Head

The classification head transforms the global context features into binary classification outputs through a multi-layer neural network architecture. The process begins with adaptive average pooling that converts the temporal features into a fixed-size representation regardless of input sequence length.

The multi-layer classifier consists of three sequential linear transformations:

- First linear layer projects the input features to 256 dimensions, followed by batch normalization and ReLU activation
- Dropout layer with rate 0.4 provides regularization to prevent overfitting
- Second linear layer reduces dimensionality to 128 features, followed by batch normalization, ReLU activation, and dropout with rate 0.3
- Final linear layer outputs a single logit for binary classification

The entire architecture is trained end-to-end using binary cross entropy loss with class weighting to handle potential class imbalance in ECG rhythm classification tasks.

### 2.3. Training Pipeline

#### 2.3.1. Optimization Strategy

The model had random weight initialization and was trained using the AdamW optimizer with differential learning rates applied to various model components. The embedding network was initialized with a learning rate of **1***×* **10**^−**5**^, while the classifier components utilized a learning rate of **1** *×***10**^−**4**^. Weight decay was set to **1** *×***10**^−**4**^ to regularize model parameters. Gradient clipping was applied with a parameter of 1.0 to prevent gradient explosion.

#### 2.3.2. Learning Rate Scheduling

A Cosine Annealing learning rate scheduler was implemented with minimum learning rate of **1** *×***10**^−**6**^.

This scheduler allows for initial 20 epochs with higher learning rates and then it decays gradually.

#### 2.3.3. Handling Class Imbalance

To mitigate the bias introduced by an uneven class distribution, we incorporate a class-weighted loss. Specifically, for binary classification we compute a weight for the positive class as

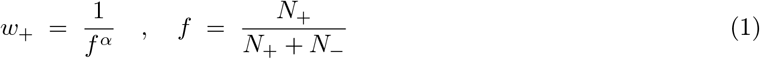

where *N*_+_ and *N*_−_ denote the numbers of positive and negative samples in the training set, respectively, *f* is the empirical positive-class frequency, and *α* is a scaling exponent (set to 1 in our experiments). The negative-class weight is taken as *w*_−_ = 1. By up-weighting the minority class in proportion to 1*/f*, the loss function emphasizes rare events and helps prevent the model from trivially predicting the majority class.

### 2.4. Experiments Performed

Various distinct experimental protocols were conducted to comprehensively evaluate model performance and generalisation. We describe the four different experiments conducted below :

1. **Multi-dataset training**: A single network was trained on the union of Code-15, Chapman, Chapman-Shaoxing, MedalCare-XL, Offline.TS and PhysioNet 2021. Evaluation was performed both on held-out internal splits and on the unseen MIMIC IV dataset. Number of training samples were 237,921, validation and testing samples were 29,740 each. Further, we tested this on MIMIC IV as a test set which includes 800,034 ECG signals. Refer table 1 to check the distribution of labels across all datasets.
2. **P-wave mask augumented training**: The most common characteristic of AFib are irregularly irregular R-R intervals and the absence of P waves. This experiment was done using a two-stage pipeline, where in the first stage we segment out P wave regions using a trained lightweight 1-D U-Net architecture. An attention module to focus on these segmentation masks was incorporated in the classifier architecture such that the model focuses on important P wave regions, which is important in diagnosis of AFib. The data set used was the same as that used in multi-dataset training with their P wave masks.
3. **Vectorcardiogram (VCG) based training**: The 12-lead ECG was transformed to 3-lead VCG signals via the Kors transformation. All the 12 lead ECG used in multi dataset training was transformed to 3 lead VCG signals. All subsequent pre-processing was done in VCG domain and classification was done using same model architecture.
4. **HDXML-only training**: The network was trained exclusively on the HDXML dataset and then tested across all remaining datasets to quantify cross-dataset generalizability from a single source. The dataset comprised of 67,432 total ECGs with 10,471 positive AFib cases and 56,961 non-AFib cases.

## 3. Experimental Results

### 3.1. Multi-Dataset Training Results

#### 3.1.1. Model Performance Metrics

We evaluated our model using standard classification metrics including precision, recall, F1-score, and area under the receiver operating characteristic curve (AUROC). The PR curve and AUC-ROC curve can be seen in (Figure 2a, 2b) respectively which illustrates the classification performance across positive and negative cases.

**Figure 2:**
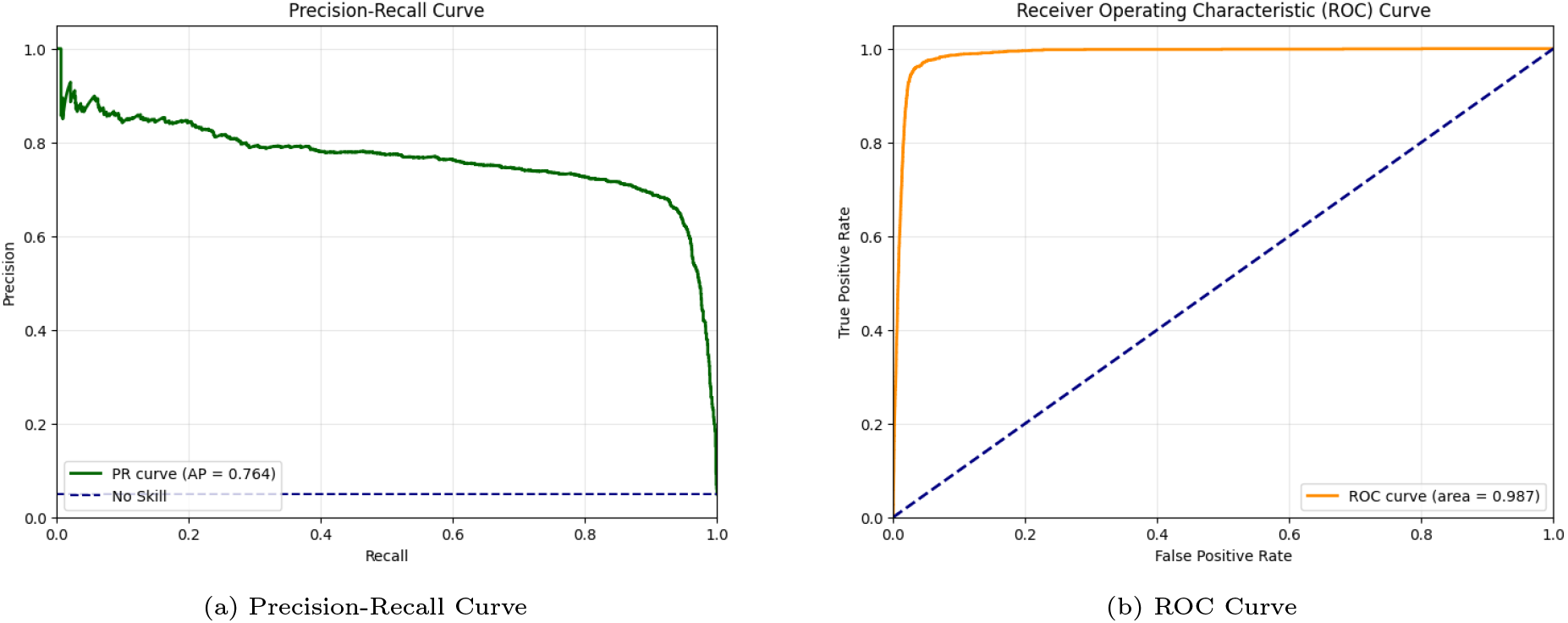
Performance evaluation metrics for multi-dataset training: (a) Precision-Recall curve demonstrating model performance across different thresholds, (b) ROC curve illustrating the trade-off between sensitivity and specificity

The model achieved an overall accuracy of 97.54% on the test set, with a precision of 84.36% and recall of 90.08% for atrial fibrillation detection. The F1-score, which represents the harmonic mean of precision and recall, was 88.50%. Sensitivity and specificity were reported to be 90.08% and 97.93% respectively. The model was evaluated with 0.7 threshold of sigmoid activation function.

The detailed performance metrics are presented in Table 2.

**Table 2:**
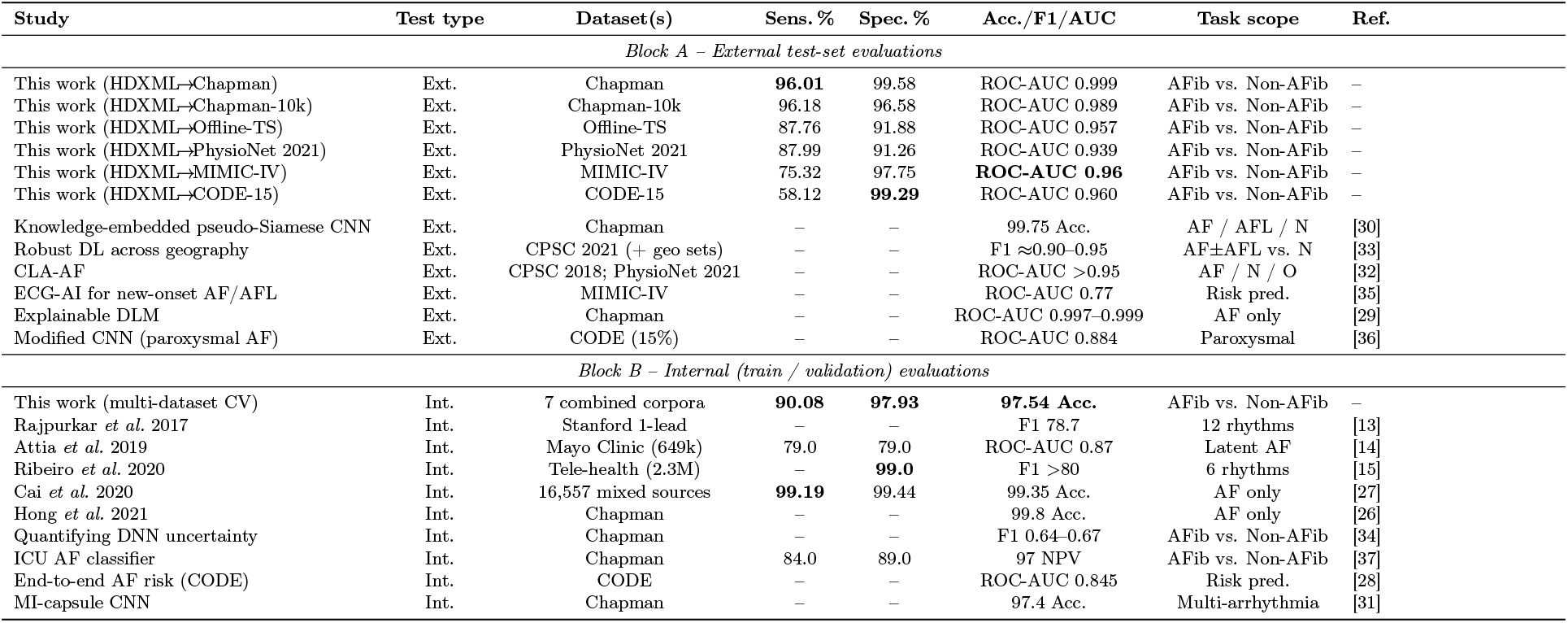
Performance of recent AF-detection systems (binary AFib vs. non-AFib unless stated). ‘Ext.” = external unseen cohort; Int.” = internal hold-out or CV. Best value in each numeric column is bold; -” = information not reported

#### 3.1.2. Evaluation on MIMIC IV ECG Dataset

The classification report revealed that the model achieved an overall accuracy of 95.71%, with a sensitivity of 70.35% and a specificity of 98.46%. The model’s precision for AFib detection was 83.26%. The F1-score was computed as 76.26%.

All evaluations were conducted on a GPU with batch processing employed to optimize memory usage and inference speed. The total execution time for the entire evaluation process was recorded as approximately 765.63 seconds, reflecting the model’s efficiency in processing large-scale ECG data.

To analyze the model’s discriminative ability across different threshold settings, a Receiver Operating Characteristic (ROC) curve was generated, and the Area Under the ROC Curve (AUC) was computed to quantify overall classification performance. A confusion matrix was computed to provide a granular breakdown of classification results, capturing true positives (correctly identified AFib cases), true negatives (correctly classified normal ECGs), false positives (normal ECGs misclassified as AFib), and false negatives (AFib cases misclassified as normal). A Precision-Recall curve was generated to assess performance in handling class imbalance, and the corresponding plot is shown in Figure 3a. Additionally, the ROC curve was visualized and can be seen in Figure 3b

**Figure 3:**
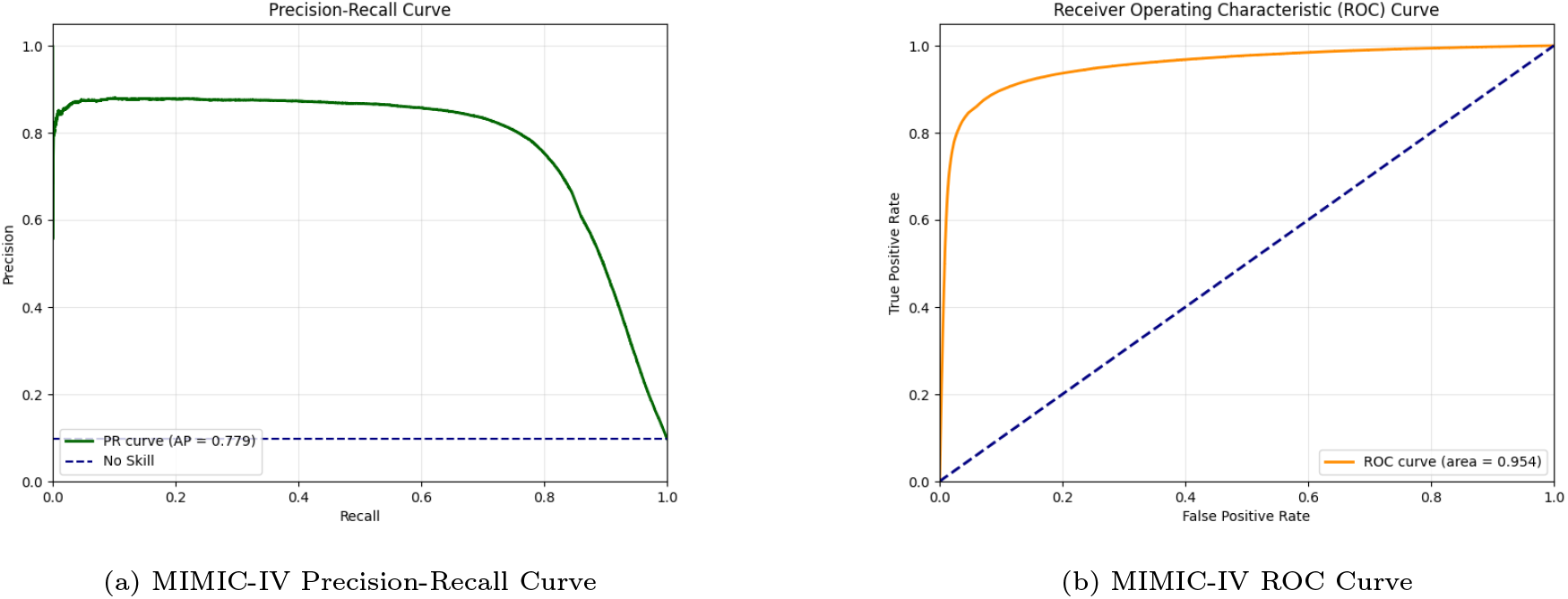
MIMIC IV external validation results: (a) Precision-Recall curve demonstrating model robustness across thresholds, (b) ROC curve illustrating discriminative performance on external dataset.

#### 3.1.3. Saliency Map Analysis

This saliency map visualization effectively demonstrates the model’s focus on clinically relevant features for atrial fibrillation detection. The low probability score, which suggests Non-AFib ECG signal, aligns with the visual evidence of regular rhythm and consistent beat morphology across leads. This approach enhances model interpretability and builds clinical trust by demonstrating appropriate attention to established diagnostic ECG features.

### 3.2. P-wave mask augumented classifier results

The ECG P-Wave AFib classifierr experiment showed variable performance across the datasets. On MIMIC IV, performance was at 95.36% accuracy with 67.80% sensitivity and 98.35% specificity.

### 3.3. VCG classifier results

The VCG-based approach using Kors transformation showed enhanced robustness to noisy signals, particularly evident in the Offline-TS dataset where accuracy improved by 3.89% and F1-score by 12.49%. However, this came at the cost of reduced sensitivity across all datasets.

### 3.4. HDXML-Only Training: Cross-Dataset Generalization

Training the model exclusively on the HDXML dataset demonstrated noteworthy cross-dataset generalization. Table 3 summarizes the performance obtained when the XML-only model was evaluated on each of the test sets.

**Table 3:** Performance of the HDXML-only model on external datasets.

| Dataset | Sensitivity | Specificity | ROC-AUC |
| --- | --- | --- | --- |
| Code-15 | 0.58 | 0.99 | 0.96 |
| Chapman | 0.96 | 0.99 | 0.99 |
| Chapman-10K | 0.96 | 0.96 | 0.98 |
| Offline-TS | 0.87 | 0.92 | 0.96 |
| PhysioNet 2021 | 0.87 | 0.91 | 0.94 |
| MIMIC-IV | 0.75 | 0.98 | 0.96 |

The results reveal four clear patterns:

1. **Strong generalization to the Chapman datasets:** Sensitivity exceeded 96% and specificity exceeded 96.5% on both Chapman test sets, indicating that features learned from HDXML transfer well to the Chapman distribution.
2. **Moderate performance on Offline-TS and PhysioNet 2021:** Sensitivity remained around 87–88% and specificity around 91–92%, showing solid but attenuated transferability.
3. **Limited generalization to Code-15:** Although specificity stayed high (*>* 99%), sensitivity dropped sharply to 58.12%, suggesting potential label noise or domain mismatch in Code-15. Despite this variability, the ROC–AUC remained above 0.93 for every dataset, underscoring the model’s strong overall discriminative capability.
4. **Interpretability Analysis** Saliency map was generated to do final class-specific analysis. Figure 4 illustrates qualitatively different attention patterns between normal rhythm and AFib cases from two of the test set samples.

**Figure 4:**
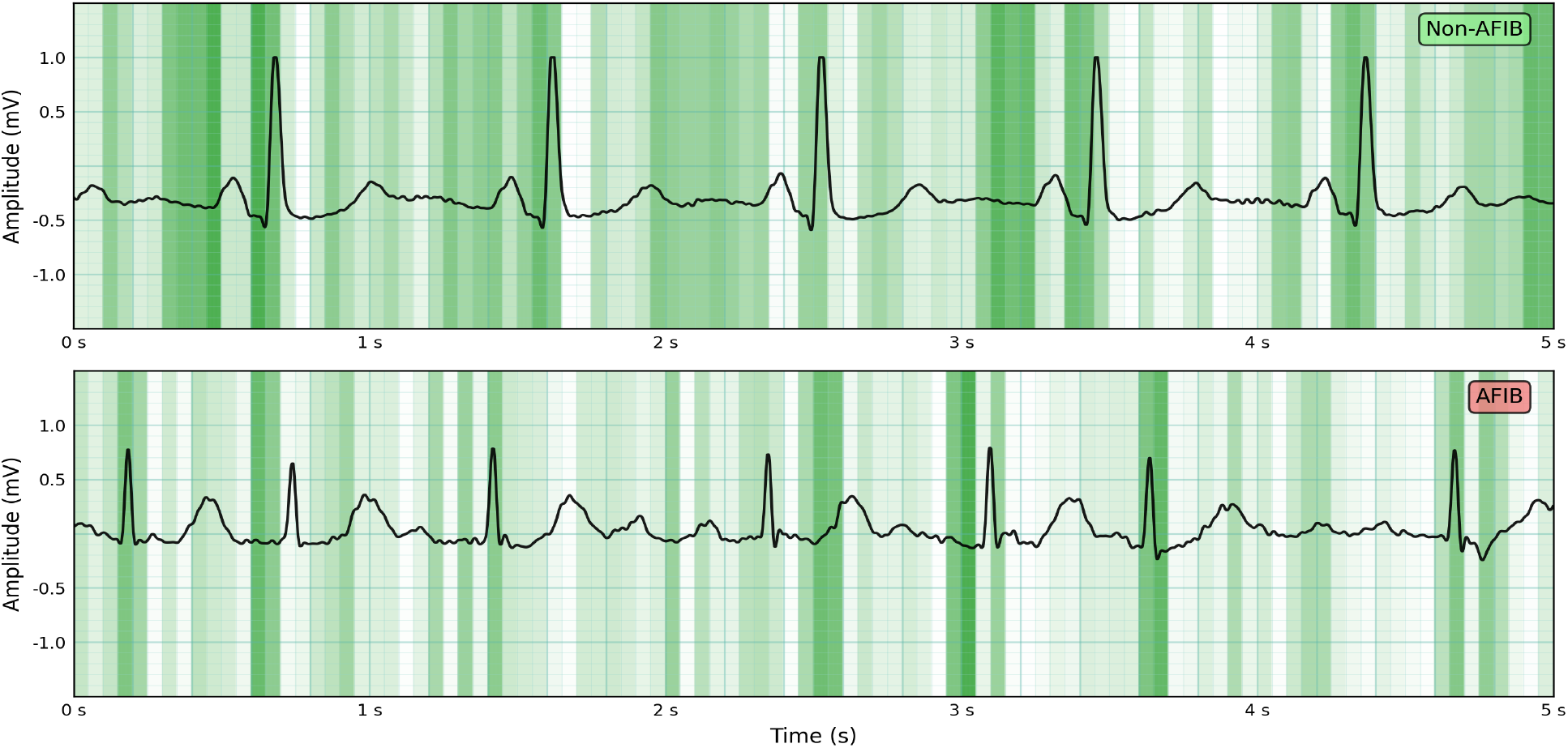
Saliency maps generated on both AFIB and non-AFIB cases.

## 4. Discussion

### 4.1. Contributions and Comparative Performance

This work addresses key gaps in the literature with the following contributions:

1. **Systematic generalisation analysis**: HDXML only experiment quantifies transferability to six unseen datasets.
2. **Deployment-oriented design**: high specificity (*>* 97%) and saliency-based interpretability meet clinical requirements.
3. **Edge-ready efficiency**: the end-to-end network contains only **17 M** trainable parameters (*≈*55MB floating point precision (FP32)) enabling real-time inference on mobile or bedside devices.

Table 2 summarises performance against nine baselines; our model offers the most balanced out-of-distribution results to date.

1. **Cross-site external validity**. Block A of Table 2 reports performance on *six* truly unseen cohorts. Competing studies typically publish a single external test (if any) and almost never include the real-world MIMIC-IV ICU records. Only ECG-AI [35] evaluated on MIMIC-IV, reporting ROC–AUC 0.77; our detector achieves **0.96**, a relative improvement of +24.7%. Likewise on Chapman, our model attains ROC–AUC 0.99 and 96.01 % sensitivity, which is slightly larger than knowledge-embedded pseudo-Siamese CNN [30].
2. **Clinical specificity under heavy covariate shift**. False positives drive alarm fatigue and unnecessary anticoagulation. Across all six external cohorts our detector keeps specificity *≥*91 %, and it remains **99.3 %** on the CODE-15 dataset. No other method in Table 2 simultaneously sustains*≥* 97 % specificity on MIMIC-IV *and* high sensitivity on Chapman Shaoxing and PhysioNet 2021 datasets.
3. **Efficiency without accuracy sacrifice**. The proposed network contains only **17M parameters** (55MB FP32), yet matches or beats heavyweight Transformers (e.g. 110 M-parameter Wide-and-Deep Transformer [17]) on every shared benchmark. This significant reduction in memory footprint enables real-time execution on mobile SoCs and edge GPUs, a prerequisite for bedside or ambulatory deployment.

Conclusively, the HDXML-only experiment demonstrates that *a single high-quality training source can yield a lightweight yet highly generalisable detector*, surpassing prior work that either lacks rigorous external validation or relies on massive, less interpretable models.

We propose this model as our final architecture, demonstrating strong potential for deployment in clinical settings due to its robust generalization and interpretability.

### 4.2. Performance Analysis

Our comprehensive evaluation demonstrates that the proposed deep learning architecture achieves state-of-the-art performance for AFib detection across diverse datasets. The HDXML-only training experiment yielded well-balanced metrics across all datasets, with the exception of CODE-15. This suggests that the CODE-15 dataset may have adversely impacted MIMIC-IV test performance in the multi-dataset training setup.

### 4.3. Clinical Relevance

Saliency map analysis confirmed that the model consistently attends to clinically meaningful regions—particularly Lead II and P-wave segments. This supports the model’s physiological alignment with AFib characteristics.

Additionally, the high specificity achieved across all settings is crucial to minimize false alarms in clinical monitoring systems, a key barrier in real-world deployment.

### 4.4. Ablation Studies and Supplementary Experiments

Several supplementary experiments were conducted en route to finalizing the HDXML-only model, which helped justify key architectural and design decisions.

#### 4.4.1. P-wave mask augumented classifier

Despite the conceptual relevance of this experiment, this approach showed inferior performance. These results were consistently lower than both the HDXML-only and multi-dataset training models. Hence, we concluded that the architecture itself effectively learns to focus on P-wave regions without the need for handcrafted segmentation.

#### 4.4.2. Vectorcardiogram (VCG)-Based AFib Detection

This approach improved robustness on certain datasets, particularly the Offline-TS set, where accuracy increased by 3.89% and F1-score by 12.49%. However, this came at the cost of decreased sensitivity across all datasets. Thus, while promising for noise resilience, VCG-based models require further refinement to match the sensitivity of multi-lead ECG models.

### 4.5. Robustness to Signal Quality

Clinical ECGs are often imperfect due to noise, lead displacements, or hardware artifacts. To simulate such variability, we introduced one random perturbation per signal per epoch during training. The perturbations included: timing shifts, lead mix-ups, missing leads, baseline drift, high-frequency noise,contact loss artifacts and smoothing.

This augmentation strategy improved the model’s ability to generalize to noisy and artifact-laden clinical ECGs, ensuring it learns robust rhythm-based representations.

### 4.6. Limitations

Performance variability across CODE 15 dataset underscores the need for careful analysis for dataset.

The model has not yet been prospectively validated in real-world clinical workflows.

## 5. Conclusion

We presented a lightweight 17M-parameter CNN that delivers state-of-the-art AFib detection from 12-lead ECGs. Two complementary training protocols were explored, yet one emerged as clearly superior:

1. **HDXML-only training is the best performer**. A model trained *exclusively* on a single, meticulously curated HDXML corpus achieved class-balanced performance on six of the six external test sets—often matching or surpassing far larger architectures—while retaining a very high mean ROC–AUC (*≥* 0.96). Only the CODE-15 cohort exhibited a marked sensitivity drop, pointing to a domain-mismatch rather than a weakness in the model itself.
2. **Real-world readiness**. The HDXML-only detector attained specificity *≥*97% on every external cohort, an essential requirement for hospital monitors and wearables where false alarms drive alarm fatigue and unnecessary anticoagulation.
3. **Clinically trustworthy and deployable**. Saliency analysis showed consistent attention to Lead II and P-wave regions, reflecting the clinical judgment of cardiologists. The small memory footprint (*≈*55MB, FP32) enables real-time inference on embedded GPUs and even high-end mobile devices.

These findings demonstrate that *one well-curated training source is sufficient to build a detector that generalizes across disparate hospitals, devices, and patient populations* with our model architecture. The HDXML-only experiment therefore offers a compelling path to regulatory approval and large-scale screening.

### Future work

We will launch a prospective emergency-department study to quantify clinical impact on alarm burden and time-to-diagnosis.In parallel, we will benchmark the network on resource-constrained edge devices—such as mobile CPUs, ARM-based single-board computers, and low-power GPUs—and explore quantization, pruning, and other compression techniques to further reduce inference latency and memory footprint.

## Data Availability

All data produced in the present work are contained in the manuscript

## Acknowledgment

The authors would like to thank the contributors of the public datasets used in this study, including the PhysioNet community, Chapman University, MIT Critical Care, HDXML for making their data available for research purposes.

